# Natural Language Processing Based Solution for Labeling Brain Metastasis Identified in Radiology Reports

**DOI:** 10.64898/2026.06.10.26355415

**Authors:** Tianhui Liu, Yu Tong Han, Hanxiao Zuo, Sunit Das, Hui-Ming Lin, Errol Colak, Marco Istasy, Aly Muhammad Ladak, Jean Claude Bigenimana, Lovedeep Gondara, Jonathan Simkin, Jaimie Lee, Daniel Roozbeh, Alan Nichol, Jacob Easaw, Emily Walker, Stephen Yip, Lili Mou, Yan Yuan

**Author notes:** co-first authors (Tianhui Liu, Yu Tong Han). equal contribution (Lili Mou, Yan Yuan).

## Abstract

**Purpose:** Brain metastases (BM) far exceed primary CNS tumours and constitute the majority workload for neuro-oncology care providers. Currently, the cancer registries only capture synchronous BMs, which is only a small proportion of all BMs. We aim to develop and validate a natural language processing (NLP) algorithm that identifies brain metastases in radiology reports, enabling scalable surveillance of asynchronous BMs.

**Methods:** Using population-based cancer registry data in Alberta, Canada, we identified a cancer cohort diagnosed between 2012–2019 with follow-up to 2022. All brain/head radiology reports at and post-cancer diagnosis were identified. Reports were sampled through a multi-phase approach and manually labeled for BM presence. We trained two Bio_ClinicalBERT models on the “Findings” and “Impressions” sections, respectively, and took the maximum predicted probability as the report-level prediction. Internal and external validation used reports from the Canadian provinces of Alberta, Ontario, and British Columbia.

**Results:** The models were trained on 1,879 samples. For internal validation, 1,833 reports from 357 patients were tested. At a probability threshold of 0.4, the model achieved a sensitivity of 0.888 and precision of 0.499. The ensemble substantially outperformed single-section models, which achieved sensitivities of only 67.8% (Findings) and 74.2% (Impressions). On external validation, sensitivity was 0.918 in Ontario and 0.726 in British Columbia, demonstrating robustness across diverse data distributions.

**Conclusions:** An NLP-based pipeline processing both Findings and Impressions sections has been developed and validated in three Canadian provinces. It meets cancer registry operational requirements and to be implemented into the surveillance workflow in Alberta and British Columbia, providing a foundation for population-level BM surveillance.

## Introduction

Brain metastases (BM) impose a severe burden on cancer patients and constitute the majority workload for neuro-oncology care providers [1, 2]. BM diagnoses far exceed those of primary brain tumors, yet accurate quantification remains elusive, as current cancer registries are not mandated to capture metastatic progression [3]. While the Canadian Cancer Registry captures the approximately 3,200 synchronous BM cases that occur annually, the burden of asynchronous BMs which develop after cancer diagnosis, far exceed synchronous BMs and are not captured in Canadian population-based databases [3, 4, 5]. Identifying these asynchronous cases would rely on manual record review by highly trained cancer registrars, which is a resource-intensive task.

Natural language processing (NLP) is a technique that can efficiently analyze vast textual data by merging artificial intelligence with linguistics. A major advance in NLP is BERT (Bidirectional Encoder Representations from Transformers), a transformer-based large language model (LLM) that uses bidirectional self-attention to learn contextual representations of text. Building on this architecture, BioBERT is a domain-specific variant of BERT further pre-trained on large-scale biomedical corpora, allowing it to better capture the specialized terminology and semantics of clinical and biomedical text [6, 7]. NLP-based approaches have already shown considerable promise in oncology report classification. For example, previous work utilized LLMs and achieved strong performance in identifying breast cancer relapse from CT reports and pathology reports [8, 9]. These studies demonstrate the feasibility of using NLP to capture outcomes that cancer registries do not routinely collect. Hence, utilizing LLMs represents a promising strategy for capturing possible BM diagnoses in the radiology reports in which they were identified.

Prior automation attempts have largely relied on keyword extraction or single-section analysis of radiology reports [10]. While recent transformer-based studies (e.g., RadBERT, GatorTron) successfully infer metastatic sites, they often restrict analysis to the “Impression/Conclusion” section [11], potentially missing up to 36% of relevant findings documented only in the “Findings” section [12]. Moreover, existing studies have rarely evaluated model generalizability across heterogeneous data sources or validated performance within real world workflow. To address these gaps, we developed an ensemble architecture that processes both the “Findings” and “Impressions” sections from CT, MRI, and PET reports (details shown in Table S1). The probability threshold is chosen to maximize sensitivity in order to meet operational standards in cancer registries. To evaluate the algorithm’s robustness across diverse data distributions and reporting standards, the algorithm was developed using radiology reports from the Alberta province, then externally validated on radiology reports from two other Canadian provinces (Ontario and British Columbia).

## Methods

### Study Population

Using the population-based cancer registry data in Alberta, Canada, we identified a cancer cohort diagnosed between 2012-2019 with follow-up to 2022. For each patient in this cohort, all radiology reports related to the brain/head from their date of cancer diagnosis (± 90 days) and onward were curated and linked. This set of reports is the underlying dataset for algorithm training.

### Training Data Set Curation

To create a labeled dataset from the underlying data set of unlabeled radiology reports, we implemented an iterative curation process to address the low prevalence of brain metastases (BM) in the source data. The model was trained and iteratively refined during Phases 1 and 2, followed by internal and external validation in Phases 3 and 4, respectively. In each phase, reports were manually annotated using a four-tier scale (“yes,” “likely yes,” “likely no,” and “no”); these were binarized into positive and negative classes for classification. The development process is illustrated in Figure 1.

**Figure 1.**
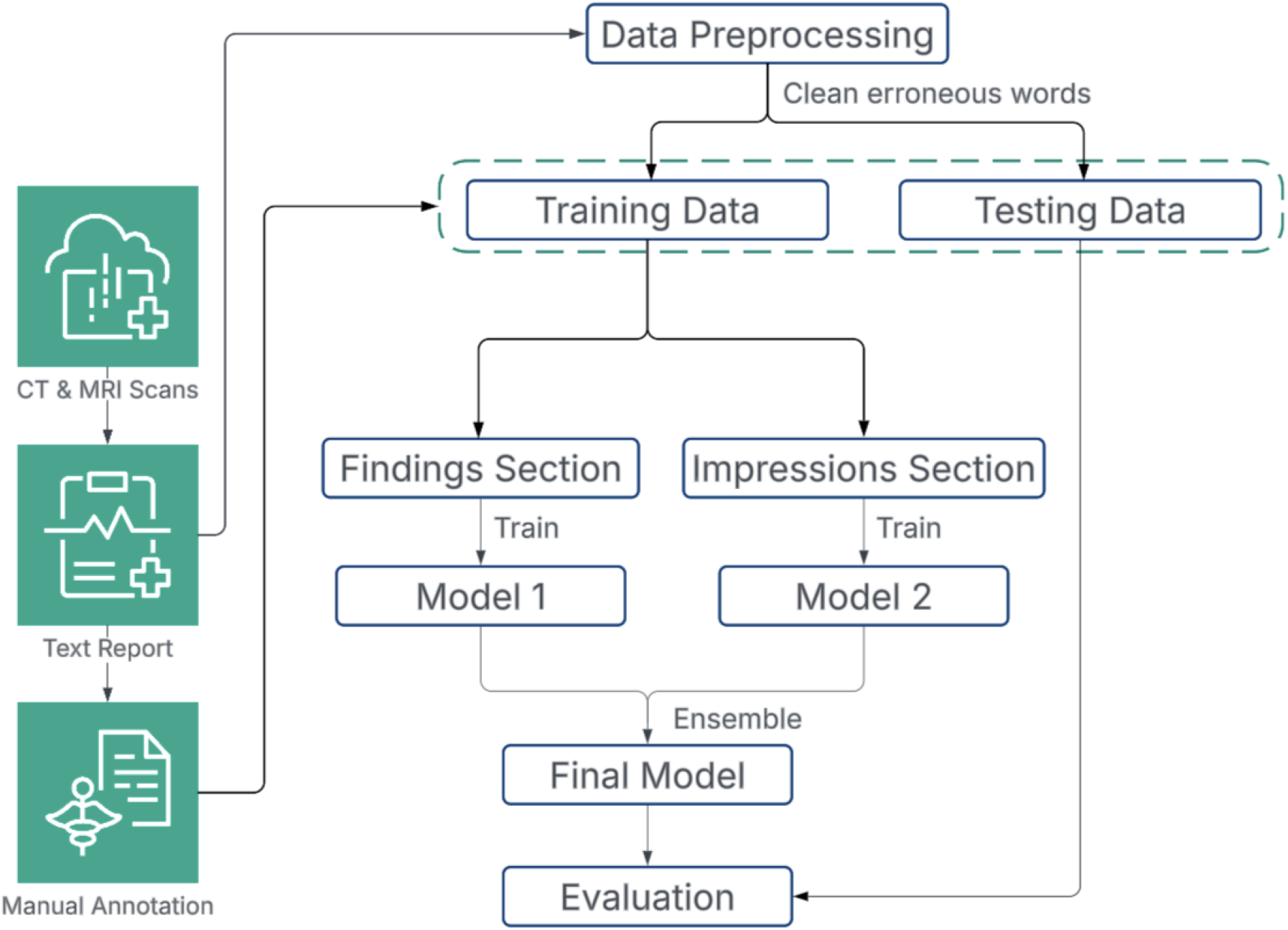
Overview of NLP pipeline development.

### Phase 1: Initial Data Curation and Model Training

The first phase aimed to identify sufficient positive BM cases for model training. Due to the low prevalence of BM, simple random sampling was deemed inefficient. Instead, we utilized the Alberta Cancer Registry (ACR) data to implement a stratified sampling strategy. By leveraging ACR data on primary cancer sites and synchronous metastasis sites, we classified patients into: 1) with primary brain tumors (PBT) diagnosis, malignant and non-malignant; 2) with synchronous BM at diagnosis; 3) none of the above. We then equally sampled patients from each category radiology and extracted reports within a 90-day window of their cancer diagnosis dates, creating a dataset enriched with patients with PBTs and synchronous BMs.

While the ACR data was used to sample patients and their reports, it was not directly used to assign report labels, as cancer registrations are derived from multiple data sources that extend beyond radiology reports (e.g. pathology reports). Consequently, an experienced research assistant under the guidance of a radiologist conducted manual annotation to assign labels based solely on the content of the radiology reports. A total of 1,200 reports were sampled and annotated for BM presence.

### Report Preprocessing and Model Training

Drawing from other relevant studies, we selected Bio_ClinialBERT – initialized with BioBERT and pre-trained on the MIMIC-III [13] database – as our base model. To address its 512-token limit, which would have truncated 25% of the full reports (Supplementary Figure S2), we isolated the “Findings” and “Impression” sections using regular expressions. Separate models were trained for each section, and their outputs were ensembled by taking the maximum predicted probability. A report was classified as BM-positive if this final probability exceeded a predefined threshold.

### Phase 2: Model-Guided Stratified Sampling and Model Training

The primary objective was to identify asynchronous BMs in post-diagnosis reports, which were excluded from the Phase 1 training set. Consequently, Phase 2 employed a model-guided sampling approach to select additional reports for annotation. We applied the Phase 1 model to 30,000 randomly selected post-diagnosis reports and used the resulting predicted probabilities for stratified sampling (Table 2). From this pool, 1,011 reports were selected for manual annotation. The final model was then trained on the combined dataset from Phases 1 and 2.

### Phase 3: Internal Validation

The internal validation cohort included patients diagnosed with a first primary cancer between October 1, 2012, and September 30, 2013, who did not have a primary brain tumor or synchronous BM (Figure 2). Radiology reports of brain/head from at least 15 days post-cancer diagnosis were included for the validation cohort which was followed up to Oct 31, 2022. There is no overlap between reports in the internal validation data and the training data.

**Figure 2.**
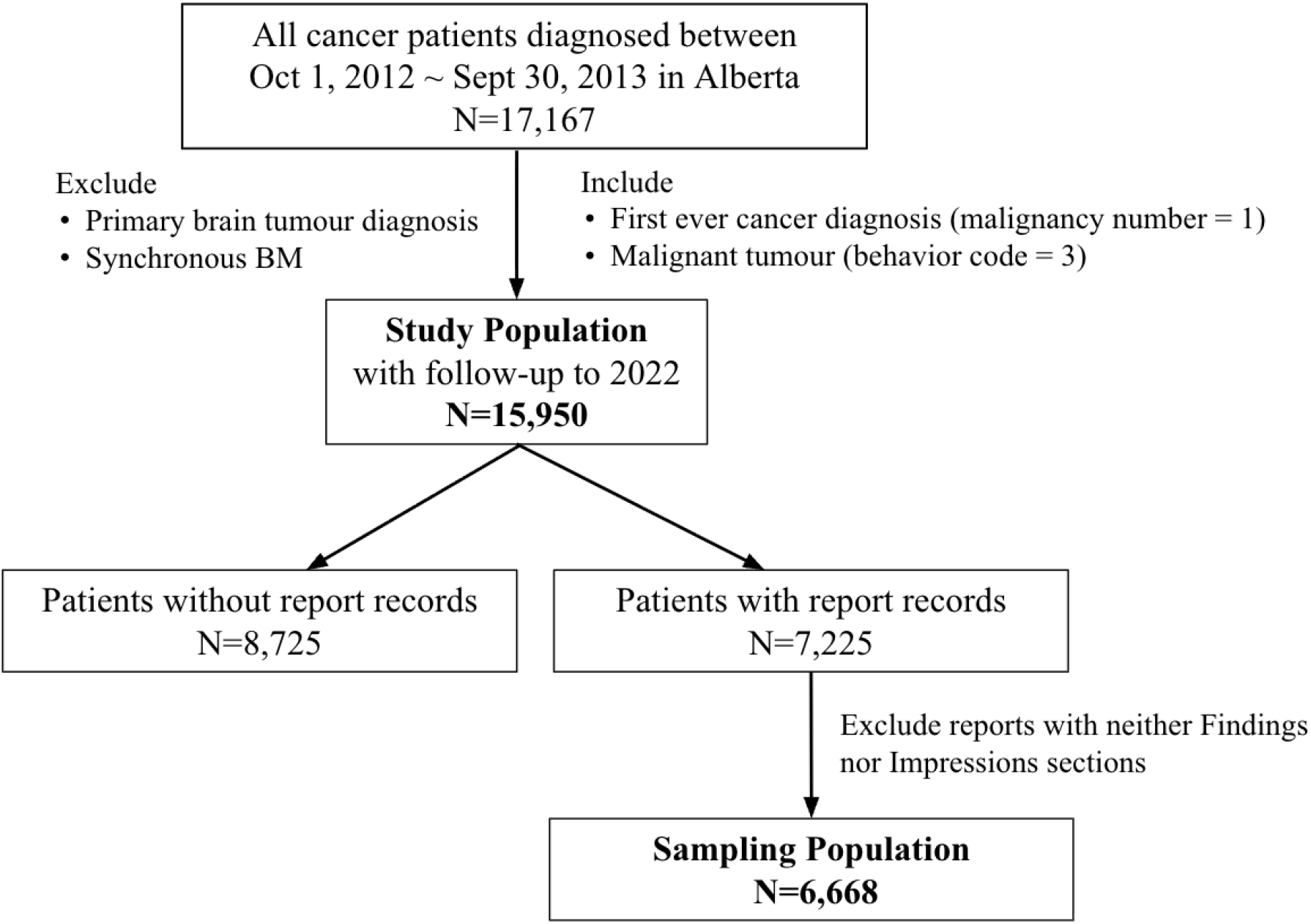
Flowchart of internal validation report sampling.

#### Missing section solution

Although our model training relied on reports containing both “Findings” and “Impressions” sections, real-world data may contain one or neither section. We restricted our internal validation dataset to include reports containing at least one of the two sections to maximize data utilization. When a section was missing (e.g., Findings), we imputed it with the full report text, which was then processed by the corresponding section-specific model. A report was classified as BM-positive if the maximum predicted probabilities of the two component models exceeded a predefined threshold.

### Phase 4: External Validation

#### Province of Ontario

Radiology reports from January 1, 2012, to December 31, 2022, were retrieved via keyword-based queries for adult patients (≥18 years) at St. Michael’s Hospital, a tertiary neuro-oncology center in Toronto, Ontario. Detailed extraction protocols are provided in Supplementary Materials, Part A. After deduplication and parsing, 17,402 unique reports containing at least one valid section were retained. A validation set was then formed through stratified sampling based on model-predicted probabilities (sampling scheme in Table S3). Two medical students (MI and AL) independently annotated the reports for BM, PBT, and malignancy, with discrepancies adjudicated by a senior neurosurgeon (SD). To ensure evaluation specifically targeted brain metastases, reports labeled as primary brain tumors were excluded from model performance evaluation.

We employed the same ensemble strategy as internal validation, where the final prediction was the maximum probability of the two component models. Performance was evaluated at a pre-selected probability threshold accounting for sampling scheme (algorithm is shown in Figure S2 in the supplementary materials).

#### Province of British Columbia

We also performed external validation using a population-based dataset from the British Columbia (BC) Cancer Registry. We retrieved 2,448 radiology reports with suspected CNS tumors between November 2024 and January 2025 (see Supplementary Materials, Part B). Following the same preprocessing and stratified sampling protocols used for the Ontario cohort, we selected a subset for manual labeling (Table S2). The final external validation set comprised 908 reports, which were independently reviewed by two annotators (JL and DR). Inter-annotator discrepancies were adjudicated by a senior investigator (YY).

For model evaluation, the handling of missing sections differed slightly from the Ontario cohort and internal validation. Whereas missing sections were imputed with full text in Ontario and Alberta, missing sections in the BC dataset were treated as empty strings. The ensemble strategy remained consistent across the three validation datasets: the final prediction was based on the maximum probability of the two component models.

## Results

The cancer diagnoses of patients in the phase 1 sample is summarized in Table 1. To ensure high data quality for model training, we included reports containing both required sections and excluded reports whose label by the annotator differ from the cancer registry’s classification. This process resulted in 868 annotated reports for the first stage of model training.

**Table 1.** Phase 1 Sample Stratified by Cancer Diagnosis.

| **Group** | **Number of Patients**<br>**(Number of Reports)** | **Number of**<br>**Sampled Patients**<br>**(Number of Reports)** |
| --- | --- | --- |
| Malignant PBT | 2,224 (2,228) | 190 (190) |
| Non-malignant PBT | 2,457 (2,493) | 202 (202) |
| BM | 2,257 (2,283) | 179 (179) |
| No BM and No PBT | 81,202 (95,798) | 297 (297) |
| **Total** | **88,140 (102,802)** | **868 (868)** |

PBT: primary brain tumour

In Phase 2, we performed model-guided stratified sampling to select 1,011 reports from a candidate pool of 30,000 reports, using predicted BM probabilities from the Phase 1 models (Table 2). Although 1,200 reports were initially sampled, those with missing sections were excluded from Phase 2 training. The final consolidated training set comprised 1,879 reports, with 505 (26.9%) identified as BM-positive. Patient characteristics of the training data for the NLP development are summarized in Table 3.

**Table 2.**
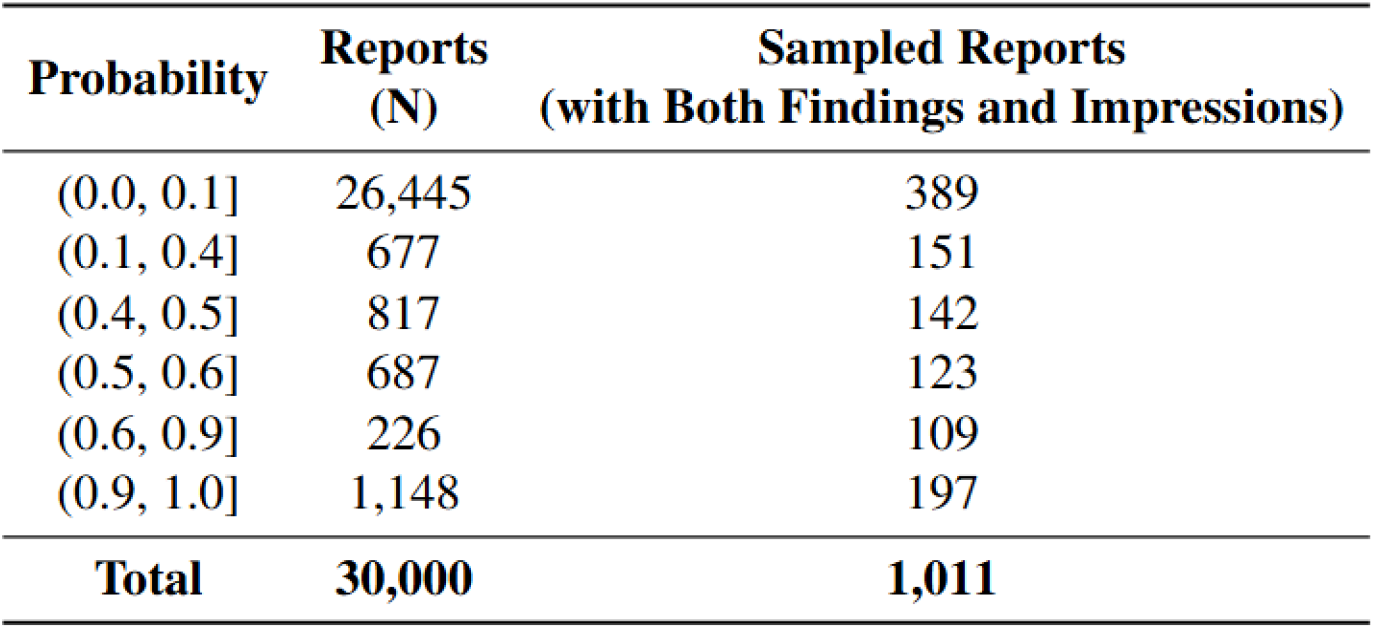
Phase 2 Sample, the Model-Guided Stratified Sampling.

**Table 3.** Characteristics of Patients Whose Reports Were Sampled in Phase 1 and 2, and Used for Model Training.

| **Characteristics, n(%)** |  | **Phase 1** | **Phase 2** |
| --- | --- | --- | --- |
| Sex | Female | 459 (52.9%) | 515 (50.9%) |
|  | Male | 409 (47.1%) | 496 (49.1%) |
| Age Group | 0–14 | 21 (2.4%) | 29 (2.9%) |
|  | 15–39 | 76 (8.8%) | 111 (11.0%) |
|  | 40–64 | 352 (40.6%) | 470 (46.5%) |
|  | ≥65 | 419 (48.3%) | 401 (40.0%) |
| Cancer Site | Brain | 200 (23.0%) | 179 (17.7%) |
|  | Bronchus/Lung | 206 (23.7%) | 219 (21.7%) |
|  | Breast | 25 (2.9%) | 89 (8.8%) |
|  | Melanoma of Skin | 13 (1.5%) | 41 (4.1%) |
|  | Colorectal | 22 (2.5%) | 44 (4.4%) |
|  | Kidney/Renal Pelvis | 15 (1.7%) | 26 (2.6%) |
|  | Esophagus | 2 (0.2%) | 7 (0.7%) |
|  | All other sites | 385 (44.3%) | 406 (40.2%) |
| BM | Positive | 179 (20.6%) | 326 (32.2%) |
|  | Negative | 689 (79.4%) | 685 (67.8%) |
| Stage | 0/I | 55 (6.3%) | 95 (9.4%) |
|  | II | 25 (2.9%) | 75 (7.4%) |
|  | III | 16 (1.8%) | 68 (6.7%) |
|  | IV | 142 (16.3%) | 179 (17.7%) |
|  | NA | 631 (72.6%) | 594 (58.8%) |
| **Total** |  | **868** | **1,011** |

The internal validation data set consists of 1,833 reports from 357 patients (Table S5 in supplementary material), of which 306 reports (16.7%) were manually labelled as BM positive and used as the ground truth. Accounting for the sampling scheme (Supplementary Figure S4), we estimated the sensitivity to be 0.888 and 0.884 for threshold probability of 0.4 and 0.5, respectively (Table 4). The corresponding precisions were estimated to be 0.499 and 0.501, respectively. We selected threshold 0.4 for external validation to prioritize sensitivity. Performance metrics for external validation are also presented in Table 4. Despite the British Columbia cohort’s significantly higher proportion of PBT compared to the training set, the model demonstrated robust generalization with a sensitivity of 0.726.

**Table 4.** Estimated Model Performance Metrics (Report Level).

| **Validation Data** | **BM Positive Report (%)** | **Threshold** | **Precision** | **Sensitivity** | **Accuracy** | **F1-Score** |
| --- | --- | --- | --- | --- | --- | --- |
| Alberta (Internal) | 11.6% | 0.4 | 0.499 | 0.888 | 0.928 | 0.639 |
|  |  | 0.5 | 0.501 | 0.884 | 0.929 | 0.640 |
| Ontario (External) | 11.1% | 0.4 | 0.739 | 0.837 | 0.843 | 0.785 |
| British Columbia (External) | 8.6% | 0.4 | 0.835 | 0.726 | 0.884 | 0.777 |

## Discussion

In this study, we developed and validated an NLP-based pipeline that identifies BM in radiology reports. By applying an ensemble of Bio_ClinicalBERT models to Findings and Impression sections, our approach identifies asynchronous BMs which are not currently captured in the cancer registry. In the internal and the external validation, the model achieved high sensitivity (0.89 in Alberta and 0.84 in Ontario) and maintained reasonable precision (0.50 in Alberta and 0.74 in Ontario) as shown in Table 4, despite substantial differences in report structure and source data.

A key strength of this work is its direct alignment with a real-world surveillance gap. By leveraging routinely collected radiology reports, the pipeline provides a scalable mechanism to capture metastatic progression at the population level, an outcome not systematically collected in cancer registries. This enables more accurate estimation of metastatic incidence, and provides critical inputs for health system planning, including forecasting demand for neuro-oncology services, radiation therapy, and supportive care. Methodologically, the multi-phase study design effectively enriches BM positive reports for model training. In addition, the dual-section ensemble approach reduced information loss compared with approaches limited to only the Impressions section.

As shown in Table S4, the ensemble approach achieved a sensitivity of 0.888, whereas models trained using only the Findings or Impression sections achieved sensitivities of 0.678 and 0.742, respectively, indicating the ability to identify the majority of BM cases. The lower estimated precision in Alberta reflects the combined effects of a sensitivity-optimized threshold and the low prevalence of brain metastases. The performance trade-off is appropriate for the intended use-case, where maximizing case ascertainment is prioritized, and false positives can be resolved through manual review by cancer registrars. Notably, the model maintained acceptable performance under external validation despite substantial heterogeneity in report structure and clinical language, indicating robustness to real-world variation. Performance in the internal cohort was stable across probability thresholds, suggesting a well-defined and robust decision boundary suitable for operational deployment.

Several limitations should be noted. First, previous works have shown that radiology reports transcribed by voice recognition tools have much higher error rate compared to those composed by radiologists [14, 15]. The transcripts may omit clinically important information or contain internal inconsistencies [16, 17]. It would be ideal if reports can be reviewed and edited by human annotators or error detection softwares before being used as training data. Second, variability in wording and report structure across provinces may reduce generalizability when models are trained on data from a single data source. Previous literature has shown “domain shifts” including differences in how regional radiologists use shorthand, describe patient demographics, and follow local imaging protocols affect the model performance on external validations [18]. Finally, our models were trained on reports containing both Findings and Impressions sections, thus performance may be reduced for reports with missing sections.

Future work could focus on improving generalizability by training on data with reports from multiple jurisdictions so that the model learns from diverse report format and writing styles. In addition, exploration of more advanced language models remains of interest; however, in our experiments, the Bio_ClinicalBERT model outperforms a larger clinical language model (GatorTron [19]), possibly due to differences in pretraining data and task suitability.

Our algorithm has been integrated into the surveillance workflow in the BC cancer registry during a preliminary test implementation. Incoming radiology reports are first processed using rule-based regular expressions to extract the Findings and Impression sections. Each section is independently tokenized and evaluated using its corresponding Bio_ClinicalBERT model that estimates the BM probability. Reports exceeding the predefined threshold are flagged for manual review to confirm true positives and exclude false positives. This automated report screening pipeline enables routine registry surveillance of asynchronous BMs.

In conclusion, our study demonstrates that transformer-based NLP models can be effectively deployed to identify brain metastases from radiology reports. Our approach addresses a key gap in the existing registry database and provides a practical foundation for the surveillance of metastatic disease.

## Data Availability

The data used in this study contain personal health information from provincial cancer registries and radiology databases and cannot be made publicly available due to privacy regulations and data sharing agreements. Requests for data access may be directed to the corresponding author and are subject to approval by the relevant data custodians and ethics boards.

## Acknowledgements of Research Support

This work is supported by the Canadian Cancer Society (CCS). We thank Alberta Health Services(AHS) for providing resources and support. We also appreciate the support of the Brain Tumour Foundation of Canada (BTFC).

Portions of this work were presented at the North American Association of Central Cancer Registries Annual Conference June 3, 2025, Hartford, U.S. and the 21st Canadian Neuro-Oncology meeting May 22-24. 2025, Vancouver, Canada (poster presentation)

## Supplementary Materials

### A. Sampling Details in Ontario

Radiology reports from Ontario were identified using keyword-based queries targeting primary and metastatic intracranial tumors. The search terms included glioblastoma, meningioma, metastatic brain tumor, metastases, astrocytoma, brain mass, glioma, oligodendroglioma, pituitary adenoma, craniopharyngioma, and lymphoma. The search was restricted to adults aged 18 years and older and to CT and MR examinations defined by the following exam codes: CT0007, CT0008, CT0009, CT0010, CT0011, CT0012, CT0013, CT0014, CT0015, CT0016, CT0017, CT0077, CT0078, CT0082, CT0083, CT0087, CT0113, CT0114, CT0119, CT0120, CT0121, CT0136, CT0137, CT0138, CT0139, CT0173, CT0269, CT0270, CT0271, CT0272, CT0273, CT0274, CT0275, CT0276, CT0277, CT0278, CT0279, CT0280, CT0281, CT0282, CT0358, CT0382, CT0383, CT0414, CT0419, MRI0001, MRI0005, MRI0006, MRI0007, MRI0008, MRI0009, MRI0010, MRI0011, MRI0012, MRI0013, MRI0195, MRI0306, MRI0364, MRI0365, MRI0366, MRI0369, MRI0660, MRI0700, MRI0701, MRI0702, MRI0703, MRI0704, MRI0705, MRI0715, MRI0716, MRI0723, MRI0728, MRI0730, MRI0731, MRI0732, MRI0779, MRI0789, MRI0892, MRI0893, MRI0894, MRI0895, MRI0896, MRI0897, MRI0907, MRI0908, MRI0915, MRI0920, MRI0922, MRI0924, MRI1085, MRI1182, MRI1368, MRI1369, MRI1370, and MRI1390. Reports were extracted for the period from January 1, 2012, to December 31, 2022, among patients seen at St. Michael’s Hospital, a major neuro-oncology centre in Toronto, Ontario.

### B. Sampling Details in British Columbia

Radiology reports from British Columbia were extracted from a population-based dataset.

The extraction covered the period from November 2024 to January 2025, for patients with a tumour site classified as brain and other nervous system. The dataset comprised 2,448 imaging reports, including computed tomography (CT; n=930), magnetic resonance imaging (MRI; n=1,497), nuclear medicine (NM; n=8), and positron emission tomography (PET; n=13). The NLP algorithm was applied to these reports to generate predicted probabilities of brain metastases (BM). To ensure we capture more BM cases, a stratified sampling approach was used based on probability bins. All reports from lower-frequency bins were included, while a fixed subsample of 500 reports was drawn from the largest bin (probability 0.0–0.1), which contained the majority of observations. The final annotated dataset comprised 908 reports.

### C. Additional Results

**Figure S1.**
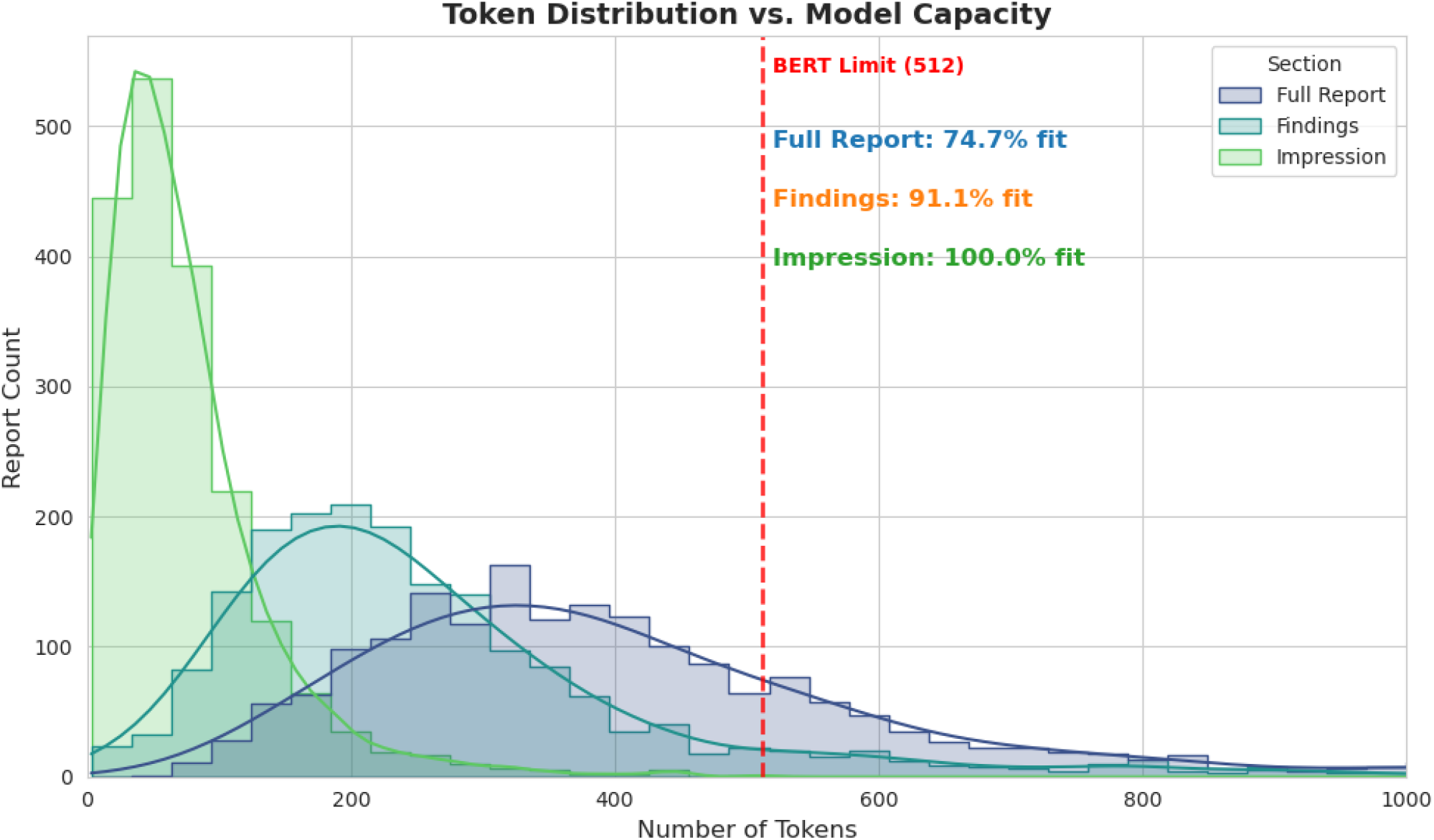
Token distribution by report sections.

**Table S1.**
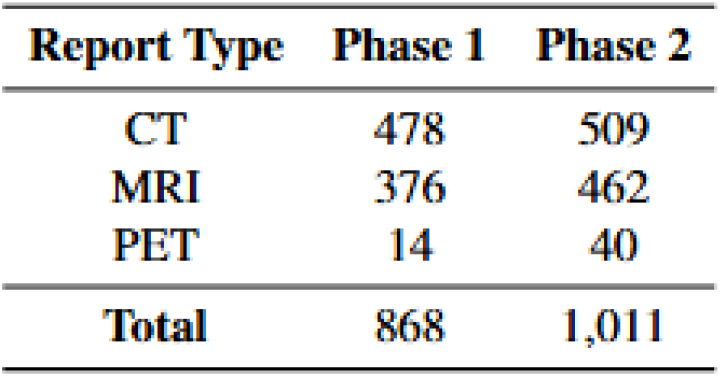
Distribution of CTs and MRIs in training data.

| Report Type | Phase 1 | Phase 2 |
| --- | --- | --- |
| CT | 478 | 509 |
| MRI | 376 | 462 |
| PET | 14 | 40 |
| **Total** | **868** | **1,011** |

**Table S2.**
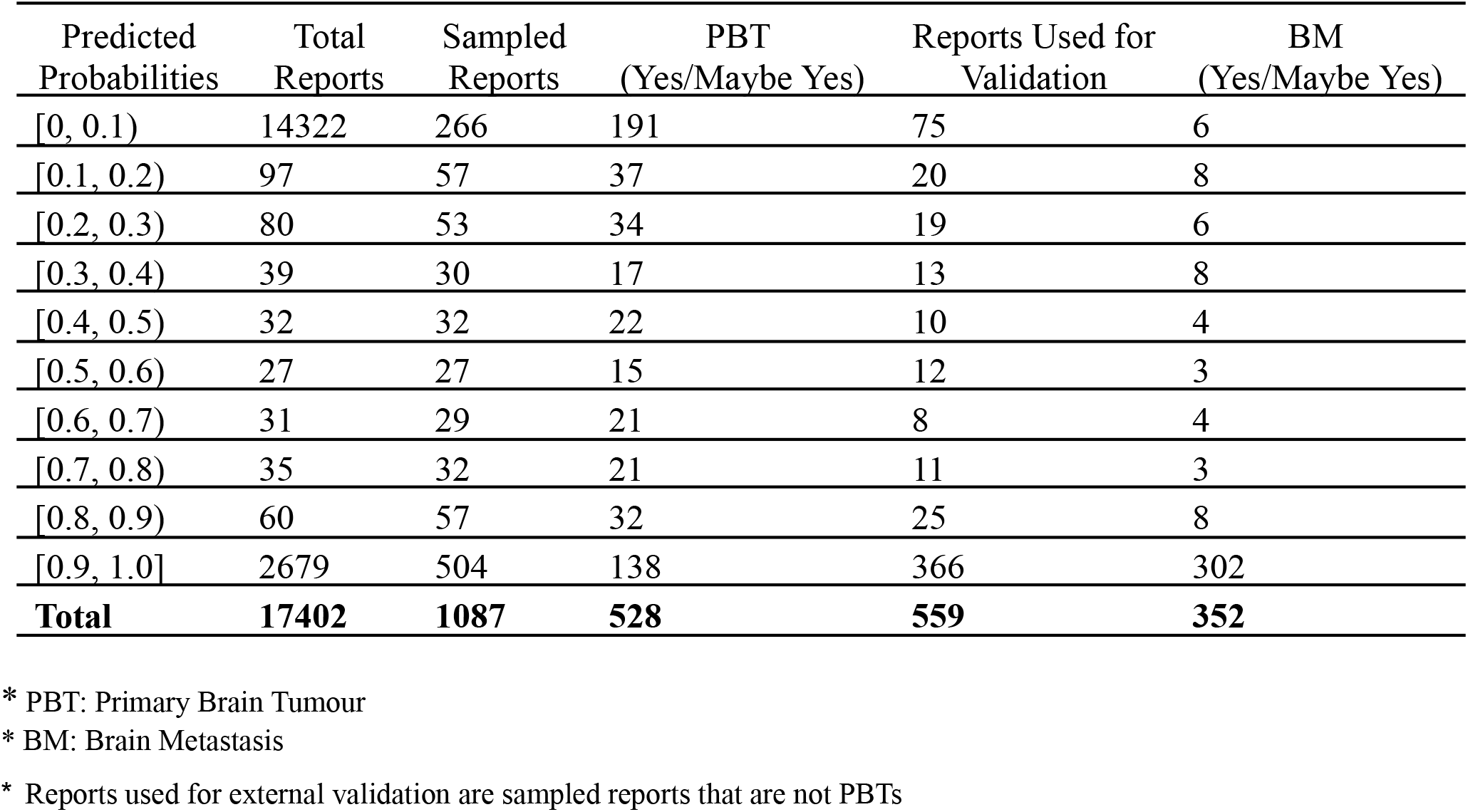
Sampling scheme for Ontario.

**Table S3.**
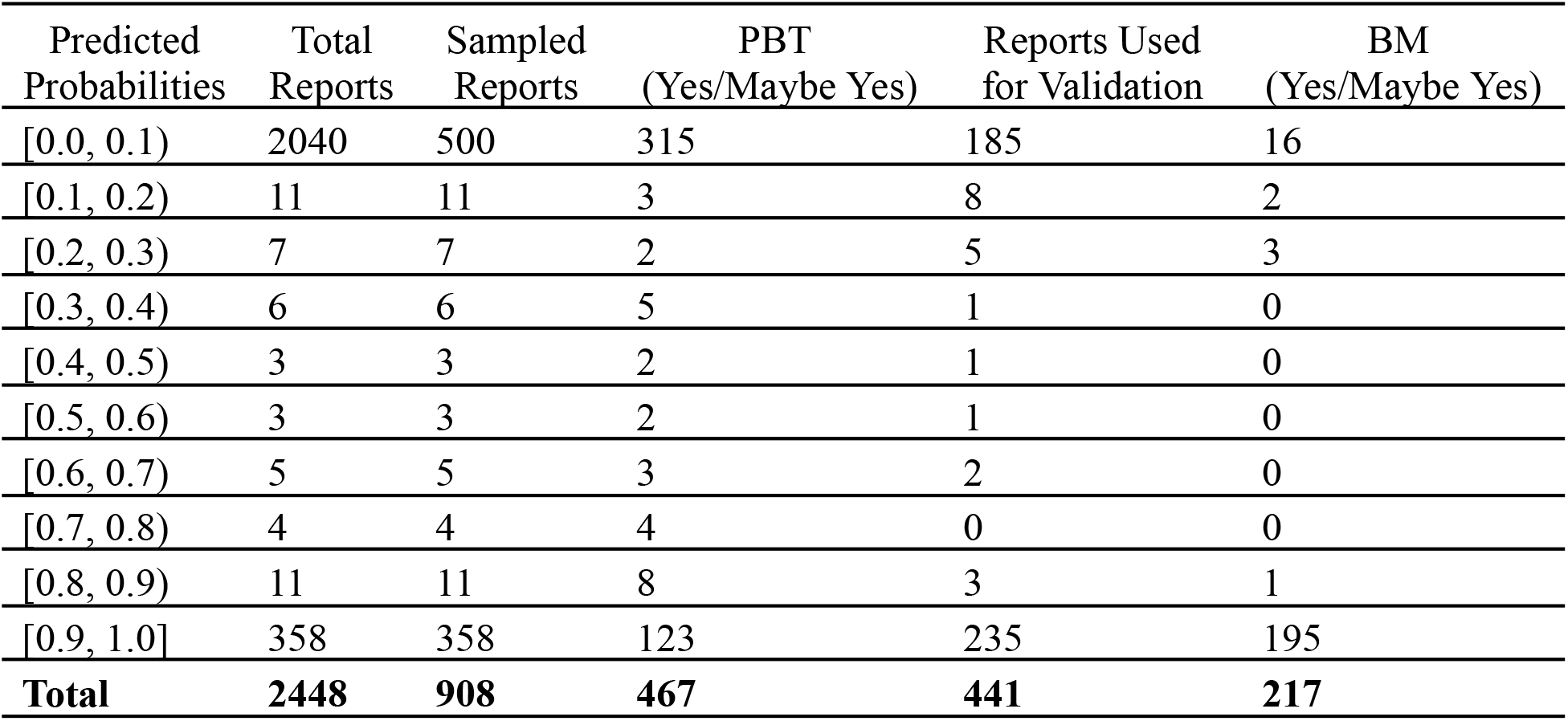
Sampling scheme for British Columnbia.

**Table S4.**
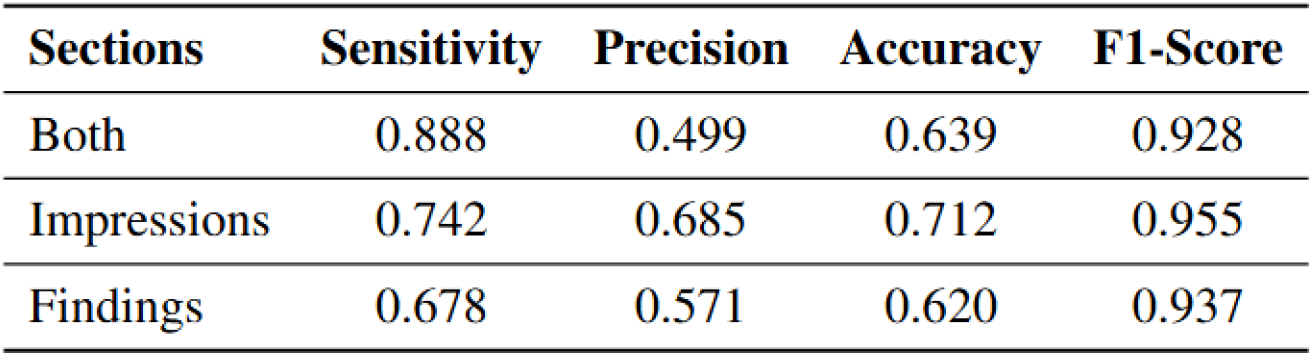
Model performance comparing the ensemble approach versus individual models trained on only one section (Internal Alberta validation dataset, threshold 0.4).

| Sections | Sensitivity | Precision | Accuracy | F1-Score |
| --- | --- | --- | --- | --- |
| Both | 0.888 | 0.499 | 0.639 | 0.928 |
| Impressions | 0.742 | 0.685 | 0.712 | 0.955 |
| Findings | 0.678 | 0.571 | 0.620 | 0.937 |

**Table S5.**
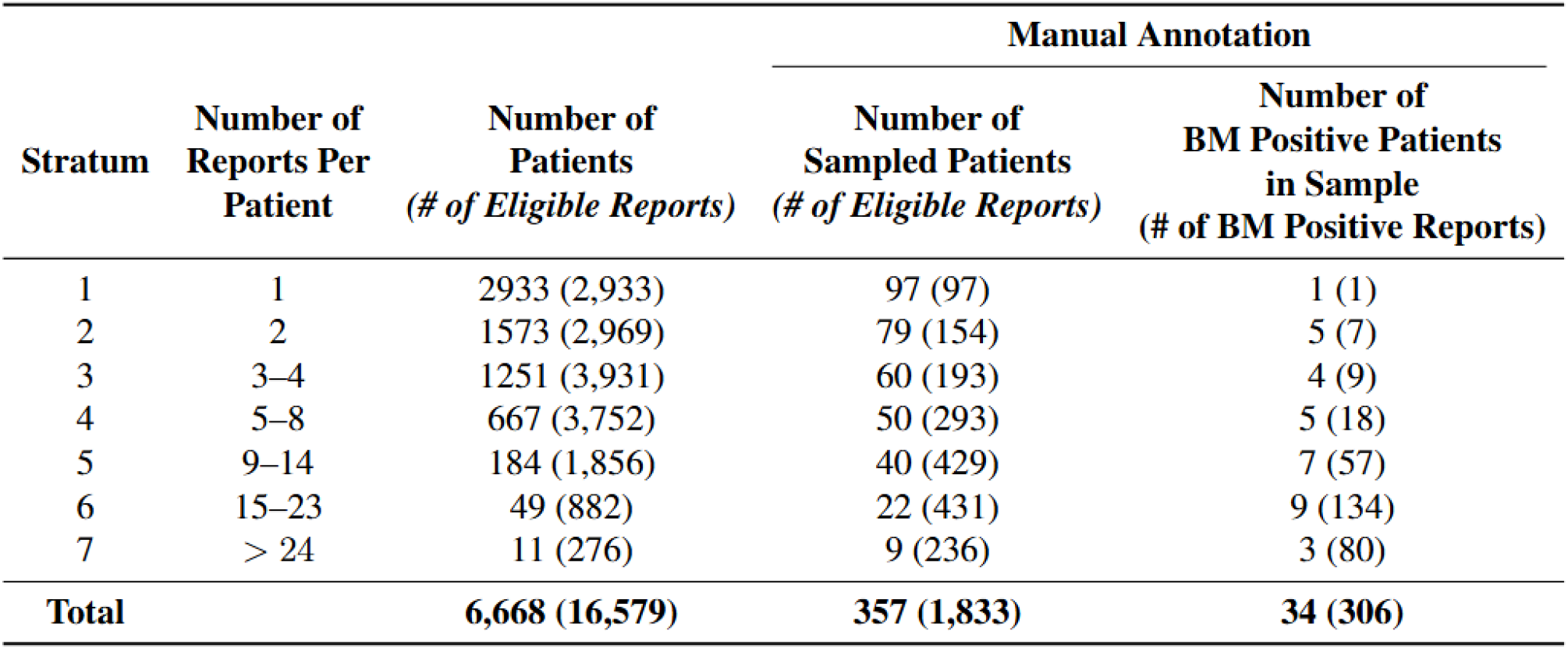
Sampling scheme of patients and reports for internal validation.

| Stratum | Number of Reports Per Patient | Number of Patients<br>(# of Eligible Reports) | Manual Annotation |  |
| --- | --- | --- | --- | --- |
|  |  |  | Number of Sampled Patients<br>(# of Eligible Reports) | Number of BM Positive Patients in Sample<br>(# of BM Positive Reports) |
| 1 | 1 | 2933 (2,933) | 97 (97) | 1 (1) |
| 2 | 2 | 1573 (2,969) | 79 (154) | 5 (7) |
| 3 | 3–4 | 1251 (3,931) | 60 (193) | 4 (9) |
| 4 | 5–8 | 667 (3,752) | 50 (293) | 5 (18) |
| 5 | 9–14 | 184 (1,856) | 40 (429) | 7 (57) |
| 6 | 15–23 | 49 (882) | 22 (431) | 9 (134) |
| 7 | > 24 | 11 (276) | 9 (236) | 3 (80) |
| **Total** |  | **6,668 (16,579)** | **357 (1,833)** | **34 (306)** |

**Figure S2.**
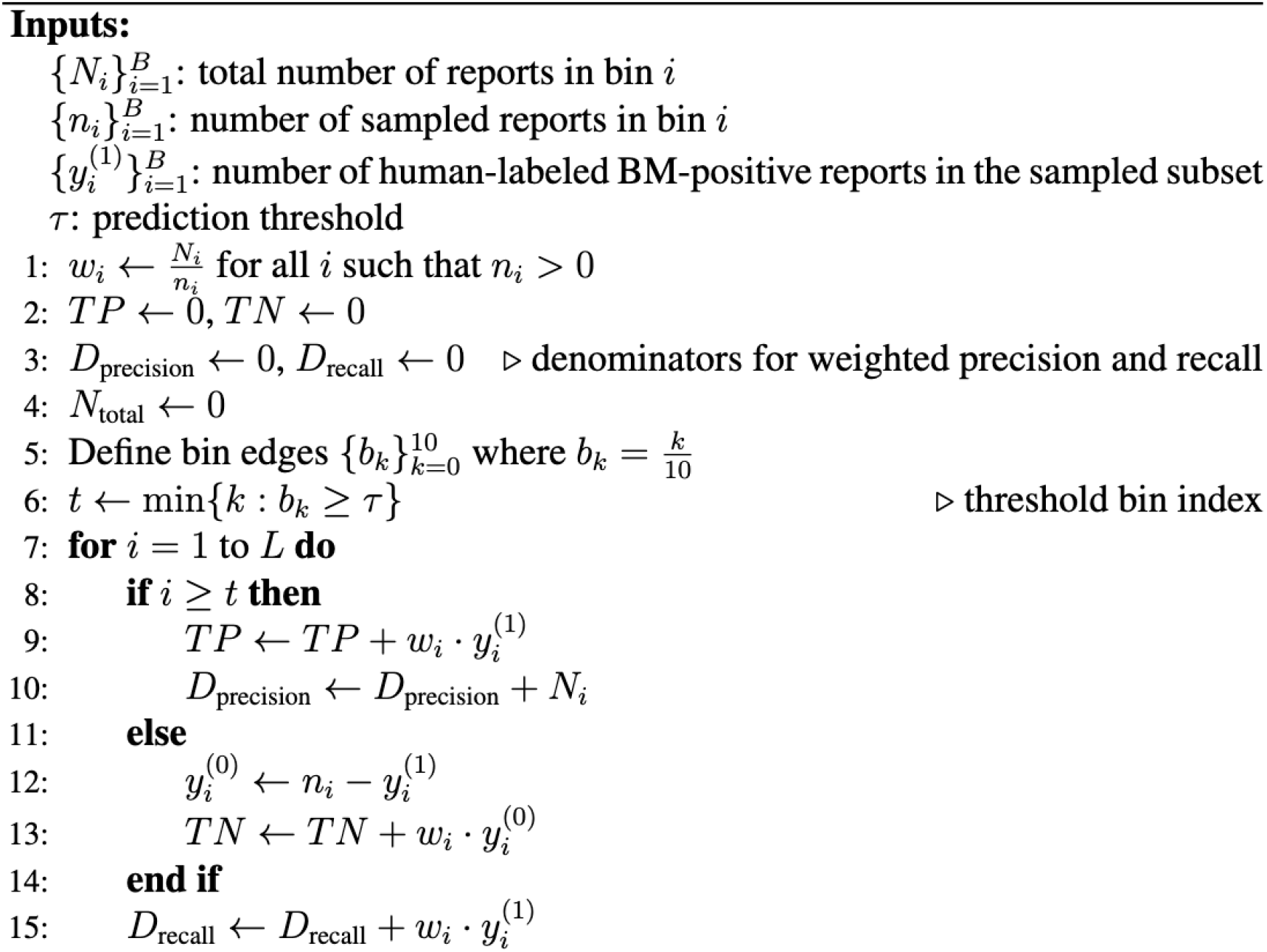
Algorithm for weighted estimates of precision, sensitivity, and accuracy from thresholded predictions for external validation, where the predicted probabilities are divided into 10 equal-width bins.

**Figure S3.**
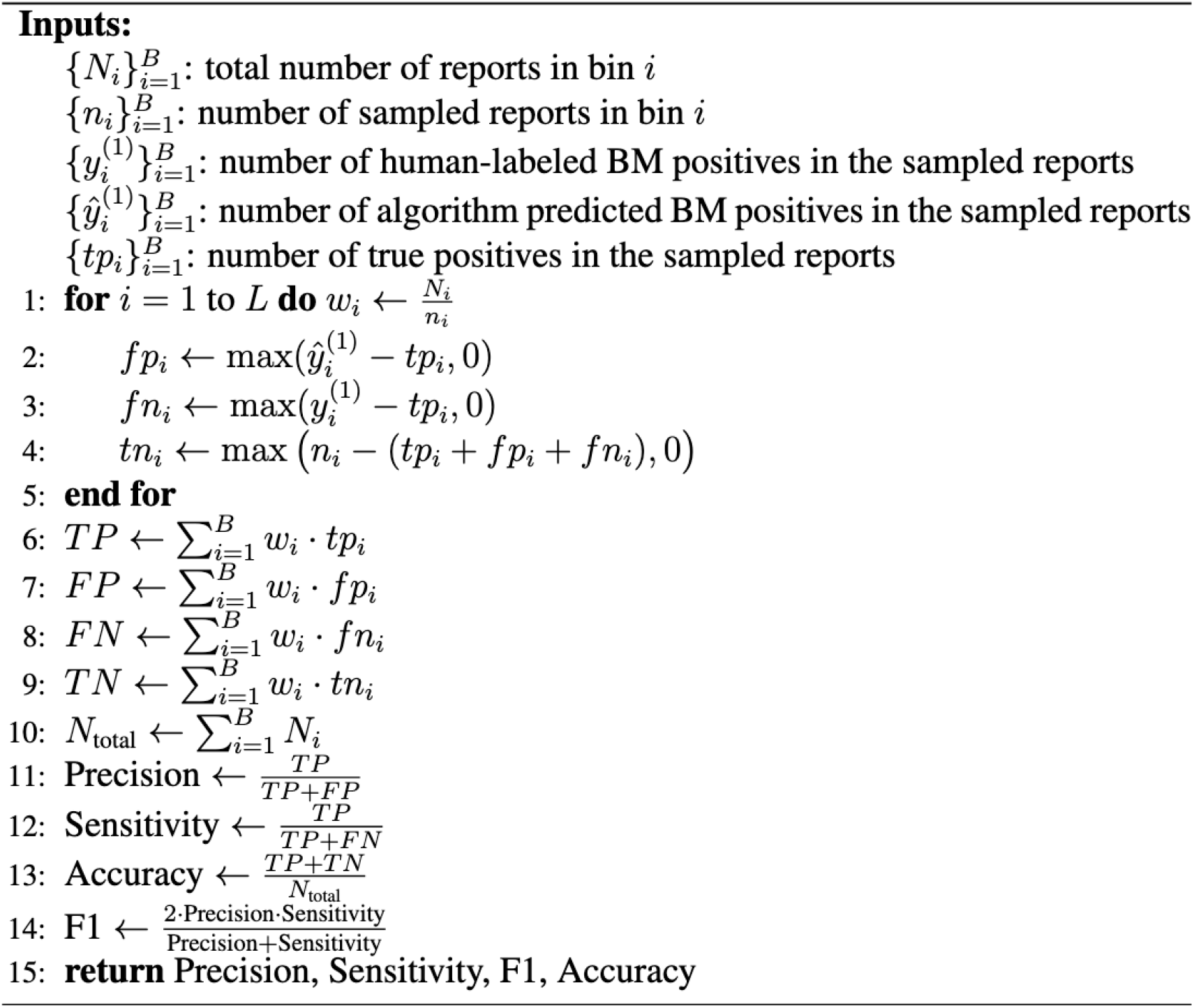
Algorithm for weighted estimates of precision, sensitivity, F1-score, and accuracy for internal validation.

## Notes

### Competing Interest Statement

The authors have declared no competing interest.

### Author Declarations

The protocol of the study is approved by the Health Research Ethics Board of Alberta, Cancer Committee (HREBA.CC-20-0179).

### Summary of Updates

We noticed discrepancy in the numbers between abstract and main text thus make a correction in the abstract. We also add a public link to our released code.

