## Supplementary Materials for "Natural Language Processing Based Solution for Labeling Brain Metastasis Identified in Radiology Reports"

C. **Additional Results**

**Figure S1. Token distribution by report sections.**

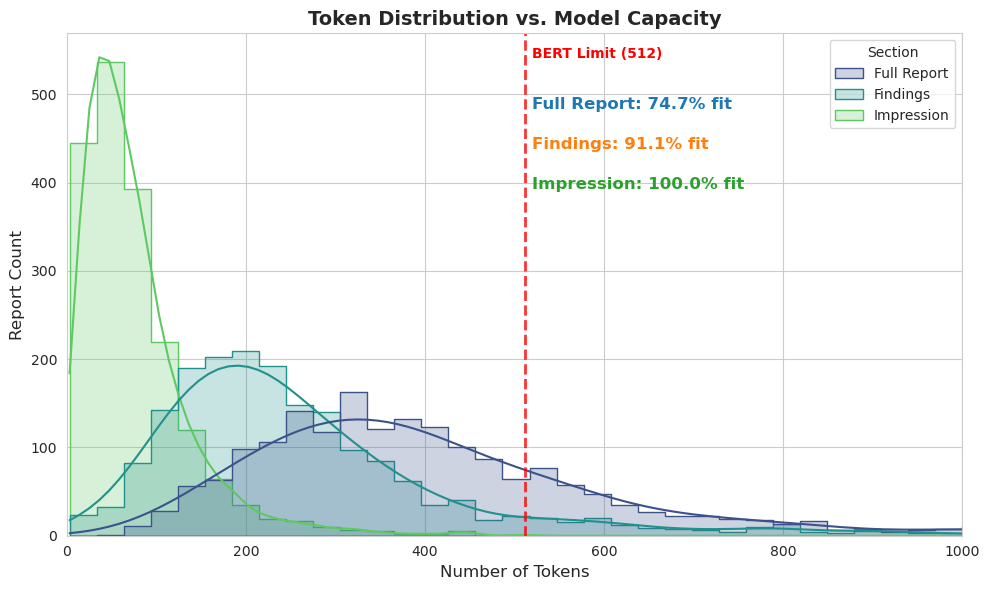

**Table S1. Distribution of CTs and MRIs in training data.**

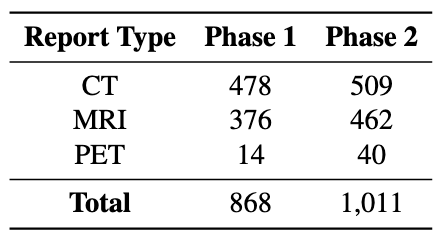

**Table S2. Sampling scheme for Ontario.**

| Predicted Probabilities | Total Reports | Sampled Reports | PBT  (Yes/Maybe Yes) | Reports Used for Validation | BM  (Yes/Maybe Yes) |
| --- | --- | --- | --- | --- | --- |
| [0, 0.1) | 14322 | 266 | 191 | 75 | 6 |
| [0.1, 0.2) | 97 | 57 | 37 | 20 | 8 |
| [0.2, 0.3) | 80 | 53 | 34 | 19 | 6 |
| [0.3, 0.4) | 39 | 30 | 17 | 13 | 8 |
| [0.4, 0.5) | 32 | 32 | 22 | 10 | 4 |
| [0.5, 0.6) | 27 | 27 | 15 | 12 | 3 |
| [0.6, 0.7) | 31 | 29 | 21 | 8 | 4 |
| [0.7, 0.8) | 35 | 32 | 21 | 11 | 3 |
| [0.8, 0.9) | 60 | 57 | 32 | 25 | 8 |
| [0.9, 1.0] | 2679 | 504 | 138 | 366 | 302 |
| **Total** | **17402** | **1087** | **528** | **559** | **352** |

* PBT: Primary Brain Tumour

* BM: Brain Metastasis

- Reports used for external validation are sampled reports that are not PBTs

**Table S3. Sampling scheme for British Columnbia.**

| Predicted Probabilities | Total Reports | Sampled Reports | PBT  (Yes/Maybe Yes) | Reports Used for Validation | BM  (Yes/Maybe Yes) |
| --- | --- | --- | --- | --- | --- |
| [0.0, 0.1) | 2040 | 500 | 315 | 185 | 16 |
| [0.1, 0.2) | 11 | 11 | 3 | 8 | 2 |
| [0.2, 0.3) | 7 | 7 | 2 | 5 | 3 |
| [0.3, 0.4) | 6 | 6 | 5 | 1 | 0 |
| [0.4, 0.5) | 3 | 3 | 2 | 1 | 0 |
| [0.5, 0.6) | 3 | 3 | 2 | 1 | 0 |
| [0.6, 0.7) | 5 | 5 | 3 | 2 | 0 |
| [0.7, 0.8) | 4 | 4 | 4 | 0 | 0 |
| [0.8, 0.9) | 11 | 11 | 8 | 3 | 1 |
| [0.9, 1.0] | 358 | 358 | 123 | 235 | 195 |
| **Total** | **2448** | **908** | **467** | **441** | **217** |

**Table S4. Model performance comparing the ensemble approach versus individual models trained on only one section (Internal Alberta validation dataset, threshold 0.4).**

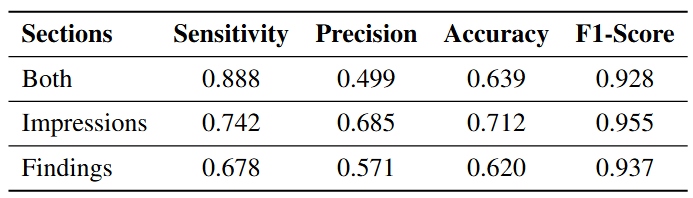

**Table S5. Sampling scheme of patients and reports for internal validation.**

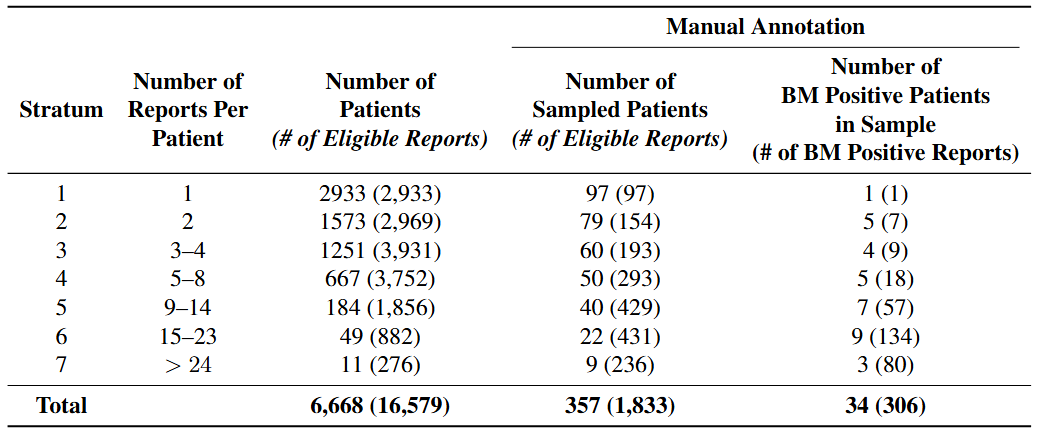

**Figure S2. Algorithm for weighted estimates of precision, sensitivity, and accuracy from thresholded predictions for external validation, where the predicted probabilities are divided into 10 equal-width bins.**

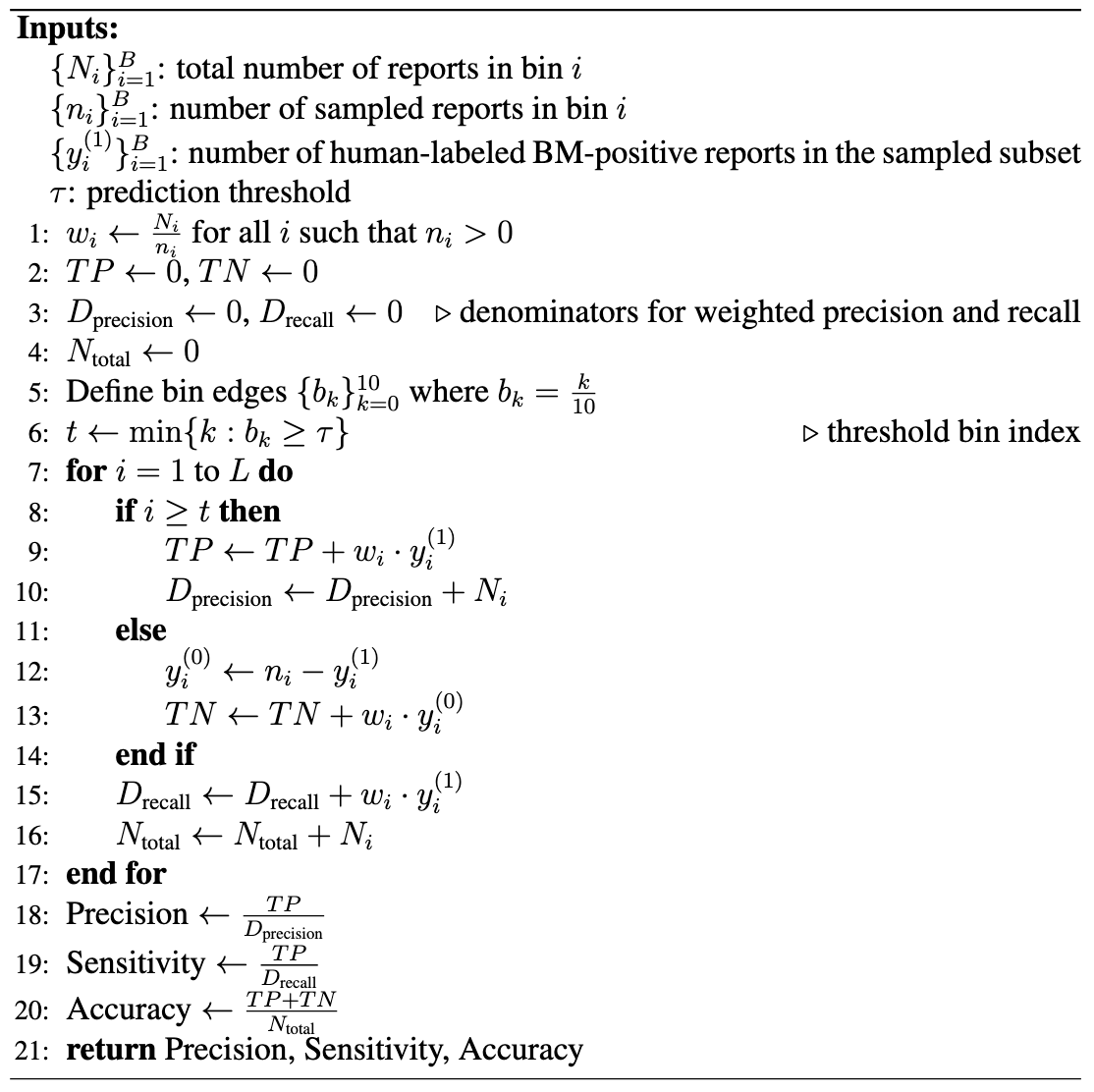

**Figure S3. Algorithm for weighted estimates of precision, sensitivity, F1-score, and accuracy for internal validation.**

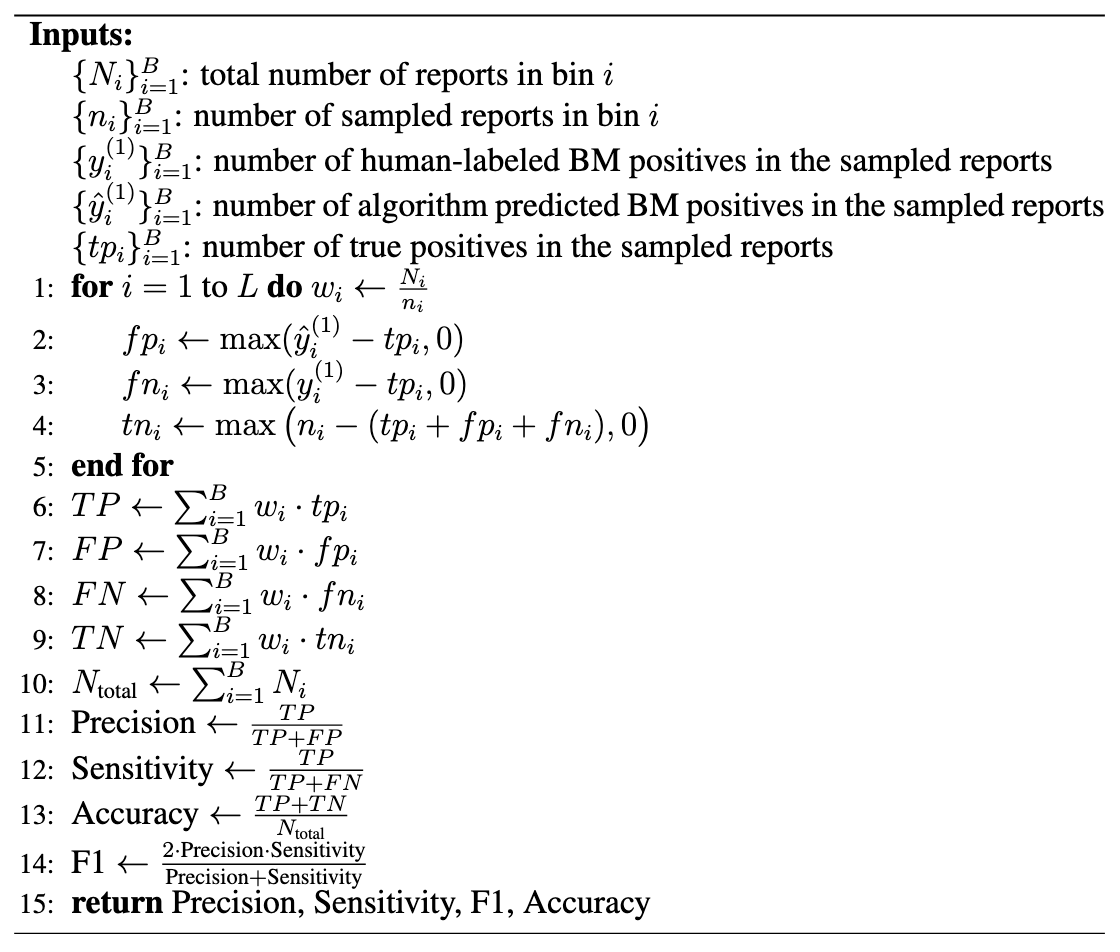
